## Supplementary File for "Trajectories of disability and influence of contextual factors among adults aging with HIV: insights from a community-based longitudinal study in Toronto, Canada"

**Supplementary Table 1**. Characteristics of participants who initiated and remained at the end of baseline monitoring phase (n = 83)

| Participant characteristics | Mean ± SD / n (%) |
|---|---|
| Age (years) | 51.5 ± 10.8 |
| Gender (n) | |
| Men | 76 (92) |
| Women | 7 (8) |
| Race/Ethnicity | |
| White | 54 (65) |
| Black or African | 5 (6) |
| Hispanic or Latino | 4 (5) |
| Number of years since HIV diagnosis | 17.9 ± 10.2 |
| Self-reported undetectable HIV viral load (<50 copies/mL) | 72 (87) |
| Current use of antiretroviral medications | 83 (100) |
| Education (some university+) | 41 (49) |
| Living alone (n) | 53 (64) |
| Number of comorbidities in addition to living with HIV (count) | 5.0 ± 4.2 |
| Three most common comorbidities | |
| Mental health condition (e.g., depression, anxiety) | 36 (43) |
| Joint pain (arthritis) | 35 (42) |
| Bone and joint disorder (e.g., osteonecrosis, osteopenia) | 31 (37) |
| Pearlin Mastery Scale score (range: 7–28) | 19.9 ± 4.0 |
| HIV Stigma Scale total score (range: 40–160) † | 93.9 ± 23.7 |
| Personalized stigma subscale score (range: 18–72) | 40.4 ± 11.6 |
| Disclosure concerns subscale score (range: 10–40) | 25.7 ± 7.0 |
| Negative self-image subscale score (range: 13–52) | 27.8 ± 8.5 |
| Public attitudes subscale score (range: 20–80) | 46.7 ± 12.6 |
| MOS Social Support Survey total score (range: 0–100) ‡ | 56.5 ± 23.8 |
| Emotional and information support raw score (range: 8–40) | 26.9 ± 7.4 |
| Tangible support raw score (range: 4–20) | 12.4 ± 5.6 |
| Affectionate support raw score (range: 3–15) | 9.5 ± 4.0 |
| Positive social interaction raw score (range: 4–20) | 13.1 ± 4.5 |

***Notes***: SD = Standard deviation. †Sixteen items on the HIV Stigma Scale belong to more than one subscale, which reflects the intercorrelations between different subscales. ‡ Raw scores of the MOS Social Support Survey were summed across the four subdomains and transformed into a 100-point scale. Greater Pearlin Mastery Scale scores, HIV Stigma Scale scores, and MOS Social Support Survey scores reflect higher levels of self-mastery, HIV-related stigma, and perceived social support, respectively.

**Supplementary Table 2**. Trajectory coefficients for physical, mental-emotional symptoms, and day-to-day activity difficulties

| | Low trajectory | | Medium trajectory | | High trajectory | |
|---|---|---|---|---|---|---|
| | Physical symptoms | | | | | |
| Model parameters | *b* | SE | *b* | SE | *b* | SE |
| Intercept | 18.01*** | 1.45 | 30.48*** | 1.98 | 42.76*** | 1.84 |
| Linear slope | -0.06 | 0.03 | -0.01 | 0.03 | -0.02 | 0.03 |
| Quadratic slope | 0.00 | 0.00 | 0.00 | 0.00 | 0.00 | 0.00 |
| | Mental-emotional symptoms | | | | | |
| Model parameters | *b* | SE | *b* | SE | *b* | SE |
| Intercept | 19.42*** | 2.13 | 35.31*** | 4.02 | 57.85*** | 2.93 |
| Linear slope | -0.08* | 0.01 | 0.02 | 0.05 | -0.02 | 0.05 |
| Quadratic slope | 0.00* | 0.00 | -0.00 | 0.00 | -0.00 | 0.00 |
| | Difficulties with day-to-day activities | | | | | |
| Model parameters | *b* | SE | *b* | SE | *b* | SE |
| Intercept | 4.98*** | 1.37 | 14.53*** | 1.37 | 35.64*** | 2.21 |
| Linear slope | -0.04 | 0.03 | 0.02 | 0.02 | -0.03 | 0.04 |
| Quadratic slope | 0.00 | 0.00 | -0.00 | 0.00 | 0.00 | 0.00 |

***Notes***: * $p$<.05. ** $p$<.01. *** $p$<.001

**Supplementary Table 3**. Trajectory coefficients for cognitive symptoms, uncertainty, and challenges to social inclusion

| | Low trajectory | | Medium-low trajectory | | Medium-high trajectory | | High/High-declining trajectory | |
|---|---|---|---|---|---|---|---|---|
| | Cognitive symptoms | | | | | | | |
| Model parameters | *b* | SE | *b* | SE | *b* | SE | *b* | SE |
| Intercept | 7.86*** | 1.94 | 18.24*** | 1.92 | 33.65*** | 2.01 | 74.21*** | 5.00 |
| Linear slope | -0.06 | 0.03 | -0.04 | 0.03 | 0.03 | 0.04 | -0.30** | 0.10 |
| Quadratic slope | 0.00 | 0.00 | 0.00 | 0.00 | -0.00 | 0.00 | 0.00* | 0.00 |
| | Uncertainty | | | | | | | |
| Model parameters | *b* | SE | *b* | SE | *b* | SE | *b* | SE |
| Intercept | 7.50* | 3.03 | 35.53*** | 1.64 | 50.26*** | 2.75 | 81.19*** | 5.06 |
| Linear slope | -0.03 | 0.05 | -0.09** | 0.03 | 0.02 | 0.04 | -0.15 | 0.09 |
| Quadratic slope | 0.00 | 0.00 | 0.00** | 0.00 | -0.00 | 0.00 | 0.00 | 0.00 |
| | Challenges to social inclusion | | | | | | | |
| Model parameters | *b* | SE | *b* | SE | *b* | SE | *b* | SE |
| Intercept | 13.46*** | 0.03 | 30.03*** | 1.43 | 40.25*** | 1.11 | 48.98*** | 1.75 |
| Linear slope | -0.12*** | 0.03 | -0.09** | 0.03 | -0.04 | 0.02 | 0.02 | 0.03 |
| Quadratic slope | 0.00*** | 0.00 | 0.00* | 0.00 | 0.00 | 0.00 | -0.00 | 0.00 |

***Notes***: * $p$<.05. ** $p$<.01. *** $p$<.001

**Supplementary Table 4**. Average posterior probability of group assignment

| Disability dimensions | Final number of trajectories | Posterior probability of group assignment | | | |
|---|---|---|---|---|---|
| | | Group 1 | Group 2 | Group 3 | Group 4 |
| Physical symptoms | 3 | 0.867 | 0.907 | 0.901 | |
| Cognitive symptoms | 4 | 0.968 | 0.847 | 0.945 | 0.920 |
| Mental-emotional symptoms | 3 | 0.939 | 0.837 | 0.862 | |
| Uncertainty | 4 | 0.983 | 0.921 | 0.885 | 0.936 |
| Difficulties with day-to-day activities | 3 | 0.897 | 0.906 | 0.966 | |
| Challenges to social inclusion | 4 | 0.943 | 0.945 | 0.905 | 0.964 |
